# COVID-19 wastewater based epidemiology: long-term monitoring of 10 WWTP in France reveals the importance of the sampling context

**DOI:** 10.1101/2021.05.06.21256751

**Authors:** A. Lazuka, C. Arnal, E. Soyeux, M. Sampson, A.-S. Lepeuple, Y. Deleuze, S. Pouradier Duteil, S. Lacroix

## Abstract

SARS-CoV-2 wastewater-based epidemiology (WBE) has been advancedas a relevant indicator of distribution of COVID-19 in communities, supporting classical testing and tracing epidemiological approaches. An extensive sampling campaign, including ten municipal wastewater treatment plants, has been conducted in different cities of France over a 20-weeks period, encompassing the second peak of COVID-19 outbreak in France. A well-recognised ultrafiltration - RNA extraction - RT-qPCR protocol was used and qualified, showing 5.5 +/-0.5% recovery yield on heat-inactivated SARS-CoV-2. Importantly the whole, solid and liquid, fraction of wastewater was used for virus concentration in this study.

Campaign results showed medium- to strong-correlation between SARS-CoV-2 WBE data and COVID-19 prevalence. To go further, WWTP inlet flow rate and raining statistical relationships were studied and taken into account for each WWTP in order to calculate contextualized SARS-CoV-2 loads. This metric presented improved correlation strengths with COVID-19 prevalence for WWTP particularly submitted and sensitive to rain. Such findings highlighted that SARS-CoV-2 WBE data ultimately require to be contextualised for relevant interpretation.

**Highlights:**

1. First study monitoring inlet of 10 WWTPs located in France for SARS-CoV-2 RNA quantification over a 20-weeks period encompassing the second peak of COVID-19 outbreak
2. Viral recovery yield was 5.5 % +/-0.5% using heat-inactivated SARS-CoV-2
3. Medium to high Spearman’s correlation strength was observed between SARS-CoV-2 WBE and COVID-19 prevalence data
4. Considering sampling context (ei. rain events) improved data consistency and correlation strength

**Graphical Abstract:** 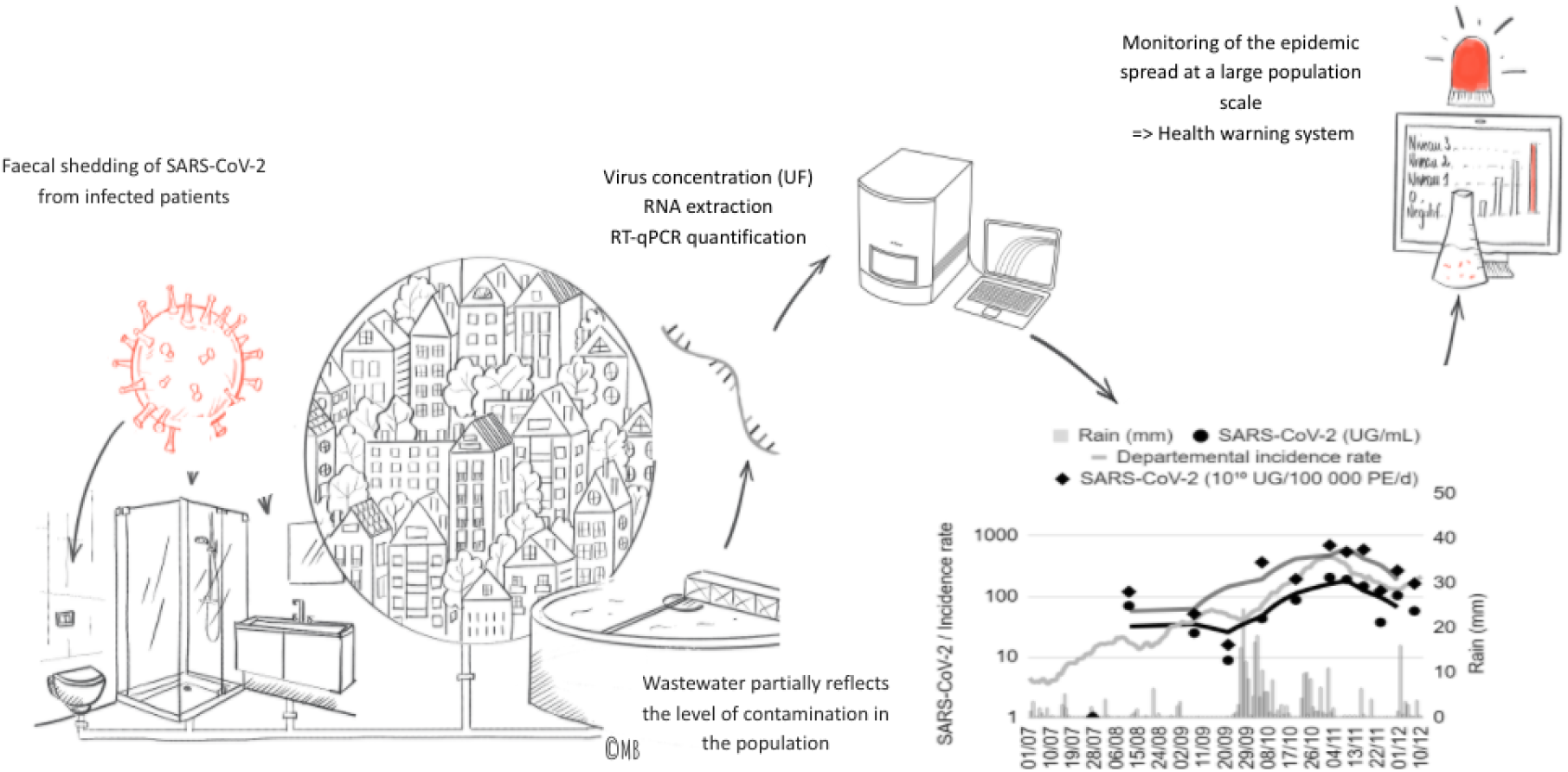

## Introduction

By March 11, 2020, WHO declared a pandemic linked to SARS-CoV-2, as 114 countries reported outbreaks of COVID-19; the first outbreak being reported in Wuhan (China) in December 2019 (WHO, 2020). France was severely affected and at the time of our last sampling (December 10, 2020), there were more than 2,130,000 confirmed COVID-19 infections and more than 57,000 deaths (Santé Publique France, 2020). As the pandemic unfolded, the water sector rapidly engaged and responded to the crisis by creating within IWA a COVID-19 Task Force whose intention is to provide the international water sector with state-of-the-art science and any measures needed to control it and monitor and protect both workers and public health. Simultaneously, the first studies investigating the relevance of SARS-CoV-2 RNA monitoring in wastewater as an indicator of distribution of COVID-19 in communities were emerging (Medema et al. 2020a, Wurtzer et al. 2020, La Rosa et al. 2020, Randazzo et al. 2020, Wu et al. 2020, Ahmed et al. 2020a, Peccia et al. 2020). Such an approach, called wastewater-based epidemiology (WBE), was theorized by Daughton (2001) for illicit-abused drugs and has been also applied to poliovirus (WHO, 2003) and other enteric viruses (Xagoraraki and O’Brien, 2020). To accelerate progress in the development of WBE applied to COVID-19 genetic signal in sewershelds, the Water Research Foundation (WRF) noted that those similar and parallel efforts should be coordinated. To address this need, WRF held the Virtual International Water Research Summit on Environmental Surveillance of COVID-19 indicators in sewersheds from 27 to 30 April 2020. During this summit, a panel of over fifty international water experts from utilities, academia, consulting and governments joined working groups and gave recommendations on best practices for sample collection, sample analysis, data interpretation and communication and appropriated quality control (Water Research Foundation, 2020). These international experts emphasized quality assurance and quality control required for sample analysis. They also stressed the need to contextualize the state of the sewershed (e.g. wet or dry weather, population connected) at the time of sampling. Only some authors take this context into account when interpreting their results. Up to now, there are not many studies on a large geographical scale combining several situations and a medium or long term follow-up (Medema et al. 2020b). There are none in France as the three published studies to date focus on 1 WasteWater Treatment Plant (WWTP) (Bertrand et al., 2020; Trottier et al., 2020) or three WWTP (Wurtzer et al., 2020). The objectives of this study were threefold: i) quantify the concentration of SARS-CoV-2 RNA at the inlet of a meaningful number of WWTP spread throughout France, ii) examine whether this monitoring is consistent with prevalence data in order to assess the relevance of WBE for COVID-19 and iii) evaluate the interest of taking into account the context of the sample collection.

## EXPERIMENTAL METHODOLOGY

Overall, our experimental procedure was set according to the recommendations of the WRF (2020), in terms of sample collection and QA/QC. Concerning sample concentration and RT-qPCR protocols, they were mainly inspired by Medema et al. (2020a and 2020b).

### Wastewater sampling

Wastewater samples were collected from ten WWTP located in France from July 27, 2020 to December 10, 2020, with a wide range of population equivalent (PE) nominal treatment capacities, from 50,000 to 555,000 PE. Composite wastewater samples (500 mL) were collected over a 24-hour period using flow-driven refrigerated auto-samplers in order to represent the average wastewater characteristics during the day. All samples were collected and kept at 4°C until RNA was extracted. All samples were analysed within a maximum of 4 days after collection.

### Samples concentration

After vortex homogenization, water samples were concentrated using 100 kDa molecular weight cutoff Amicon**™** Ultra 15 mL centrifugal devices (Merck Millipore). 30 mL of wastewater were concentrated by filtering successively twice 15 mL in the centrifugal device. Samples were centrifuged at 2,000 xg for 15 mn. After the second centrifugation, the volume in the upper part of the filter was adjusted to 600 μL with 10 mM Phosphate-Buffered Saline buffer and then homogenized 30 seconds with a vortex. Remaining biomass was removed from the filter by pipetting up and down.

### RNA extraction

RNA extraction was systematically carried out in triplicates from the 200 µL of the concentrate previously obtained. This step was performed using the Allprep PowerViral DNA/RNA kit (Qiagen), in accordance with the supplier’s recommendations, with the exception of the sample disruption step which was performed using FastPrep-24™ Instrument (MP Biomedicals™) with 2 mL Lysing Matrix tubes (MP Biomedicals™).

RNA extracts were quantified using the Qubit fluorometer (Invitrogen™) associated with the Qubit™ RNA BR assay kit, and stored at −80°C until RT-qPCR analyses.

### Quantification of SARS-CoV-2 RNA by RT-qPCR

For SARS-CoV-2 RNA quantification by RT-qPCR, two different regions of the nucleocapsid gene were targeted. N1 and N2 were amplified using primers and probes as well as PCR conditions and cycling as described in the diagnostic panel of US CDC (2020). RT-qPCR quantifications were performed using the qScript XLT One-Step RT-qPCR ToughMix (Quantabio) on CFX96 touch Real-Time PCR Detection System. The RT-qPCR reactions were carried out in simplex in order to have maximum sensitivity of the measurement. N1 and N2 probes were both labelled at the 5’-end with the FAM reporter and with the BHQ-1 as quencher at the 3’-end.

For each sample, RT-qPCR analysis was systematically performed on 2 of the 3 extraction replicates-selected based on RNA concentrations similarity-, and for each extraction replicate, the RT-qPCR reaction was performed in duplicate, i.e. a total of 4 replicates per sampling point.

Prior to the RT-qPCR, extracted RNAs were diluted with RNase free water to bring the initial RNA amount back in a range of [50-150 ng] RNA quantity per reaction. In the event that the RNA concentrations were too low to reach this level of quantity, they were by default diluted by a factor of 2, in order to remove matrix-related inhibitors. An inhibition control of the RT-qPCR reaction was systematically performed for each extraction replicate. This control consisted of a dosed addition (approximately 4000 copies/reaction) of the positive control EDX SARS-CoV-2 (Exact Diagnostic, supplied by Bio Rad) for each dilution analyzed. The amplification was considered as free of inhibition with the Cq 24 with a tolerance of 2 Cq. A standard curve, from 1.10^6^ copies/μL to 1 copy/μL, was drawn for every analysed plate using a control plasmid, N-nCoV-control-Plasmid (Eurofins). Standard curves were performed by dilution in 1/10 series with 1X Tris - EDTA. Negative control was achieved on every analyzed plate, using SARS-CoV-2 Negative Run Control (Exact Diagnostic, supplied by Bio Rad). This control is formulated in a synthetic matrix and contains 75,000 copies/mL of human genomic DNA.

For each primers/probe system, SARS-CoV-2 RNA quantification was obtained from extraction duplicate analysed in RT-qPCR duplicate each, producing 4 values by biomarker. To be conservative, the maximum among the 4 generated values was considered for further analyses.

### Data collection

COVID-19 epidemiological data were collected from the open data portal of the French National Health Agency. Precisely, 2 datasets were used: SI-DEP which stores prevalence (https://www.data.gouv.fr/fr/datasets/taux-dincidence-de-lepidemie-de-covid-19/), and global hospitalisation and death dataset (https://www.data.gouv.fr/fr/datasets/donnees-relatives-a-lepidemie-de-covid-19-en-france-vue-densemble/#_). COVID-19 incidence rate considered in this study was 7-days moving sum to compensate for delay in data collection. This weekly indicator was directly retrieved from the dataset.

The rainfall data were obtained from The Weather Company.

Operational data, such as organic loads and flow rate, were obtained directly from the ten WWTPs.

### Calculation and statistics

Population equivalents (PE) were calculated as defined in article 2 of the Council Directive 91/271/EEC of May 21, 1991 concerning urban waste-water treatment “One population equivalent means the organic biodegradable load having a five-day biochemical oxygen demand (BOD5) of 60 g of oxygen per day”.

WWTP inlet flow rate variation and rainfall correlation were evaluated using 7-days right-trailing moving average variables. Furthermore, the flow rate variation variable was computed as the percentage of flow rate variation against mean flow rate variation in 7-days averaged dry-weather conditions.

Contextualized SARS-CoV-2 RNA quantification was calculated as raw quantification impacted by the ratio of daily flow rate and averaged dry-weather flow rate.

Contextualized SARS-CoV-2 RNA load per capita was calculated as contextualized SARS-CoV-2 RNA quantification multiplied by daily flow rate and divided by PE.

Nonparametric T tests were performed to assess statistical significance between groups of data. Two-tailed p-values < 0.05 were considered significant.

Spearman’s rank correlation and Pearson’s linear regression were carried out on variables using R Tidyverse packages and p-values were checked for confidence with a 0.05 threshold p-value. For linear regression residuals patterns were checked for normality and absence of pattern for homoscedasticity.

## Results and discussion

Our first goal was to assess SARS-CoV-2 RNA quantification method performance, as recommended by WRF experts. This qualification is likely of crucial importance, as it is documented that many steps in the method can induce variability, *i*.*e*. wastewater concentration, RNA extraction, biomarker selection. In this study we will focus on ultrafiltration (UF) wastewater concentration, as it is a well-established method for SARS-CoV-2 WBE (Medema et al., 2020, Bertrand et al., 2021, Forest et al., 2021, Hemalatha et al., 2021). First results will deal with recovery yield obtained using heat-inactivated SARS-CoV-2 as surrogate. Global WWTP sample process (concentration, RNA extraction and quantification) will be qualified.

Also, concentration methods are mostly preceded by a pre-clarification step (centrifugation) aiming in solids and related PCR inhibitors removal. But it is not clear if and how this preliminary step impacts the overall quantitative SARS-CoV-2 RNA measure. Thus, we also attempt to assess the need of pre-clarification in this study.

The experimental protocol being qualified, SARS-CoV-2 WBE results obtained in this study on 10 WWTPs will be then analysed against a classical biomarker response but also considering sampling context.

### Heat-inactivated SARS-CoV-2 recovery yield

SARS-CoV-2 recovery yield was evaluated on the global analytical process, *i*.*e*. ultrafiltration, RNA extraction and N1-amplification.

To this end, a SARS-CoV-2 negative sample from WWTP1 (thereafter named NEG) sampled on August, 17 2020 was spiked with a known quantity of heat-inactivated SARS-CoV-2 (ATCC VR-1986HK™, thereafter referred as POS). POS was added into NEG with a volume ratio of 10 µL for 60 mL, producing a sample thereafter referred to as POS+NEG. After appropriate homogenization, the 60 mL POS+NEG sample was divided in two 30 mL sub-samples (1 &2) that were processed in parallel for concentration, extraction and N1-amplification steps, as described in the material and methods paragraph.

As the ATCC certificate of analysis guaranteed 3.75 × 10^5^ genome copies/μL in POS, maximum N1 concentration in POS+NEG (after the whole sample process) was 6.25 × 10^3^ genome copies/µL, constituting the theoretical 100% N1-recovery yield.

The experimental setup produced eight values evaluating not only N1 recovery yield, but also concentration and extraction repeatability.

As shown Figure 1, N1 was detected in all eight experiments, but the global recovery for ultrafiltration followed by RNA extraction was rather low with 5.5 +/-0.5% [4.8 - 6.2] of theoretical yield. Compared with other studies, such low values were already reported. For example, La Rosa et al. (2020) and Graham et al. (2020) determined recovery efficiencies of 2% and 0.1 to 7% using alternative PEG precipitation protocols with human coronavirus HCoV 229E and murine hepatitis virus, respectively. In another study, Gonzalez et al. (2020) assessed two enveloped viruses’ recovery yields -bovine coronavirus (BCoV) and bovine respiratory syncytial virus (BRSV)- with two concentration methods: ultrafiltration InnovaPrep Concentrating Pipette Select and electronegative filtration. For BCoV, total recoveries were 5.5% (± 2.1%) and 4.8% (± 2.8%) with InnovaPrep and electronegative filtration method, respectively. The recovery values reported so far vary greatly (3.3-73%) and currently, there is no consensus on the threshold recovery yield (Michael-Kordatou et al., 2020). Indeed, as highlighted by Hong et al. (2021), recovery of viral particles from untreated wastewater has been challenging as evidenced from the wide range of reported efficiencies. The reported differences in recovery efficiency can be due to the varying type of viral surrogates spiked (Hong et al., 2021). One of the most complete studies to date evaluating the impact of concentration method on viral recovery was performed on murine hepatitis virus as enveloped virus surrogate (Ahmed et al., 2020b) and reported an average recovery efficiency of 65.7% ± 23.8%. It is unclear if and how these results can be compared with heat-inactivated SARS-CoV-2 spiking. Recently, Pérez-Cataluña et al. (2021) reported high recovery yield on gamma-inactivated SARS-CoV-2, with up to 52.8 ± 18.2% for PEG-precipitation and automatic magnetic-beads RNA extraction. These data were obtained with an inactivated SARS-CoV2 virus (as in the present study) but with different concentrations and RNA extraction methods. On our side, the RNA extraction method using magnetic beads was also recently tested on our samples concentrated by ultrafiltration. Preliminary results showed average yields of 26% (data not shown). The change in extraction method appeared to improve recovery and gave yields close to those observed by Pérez-Cataluña et al 2021.

**Figure 1:**
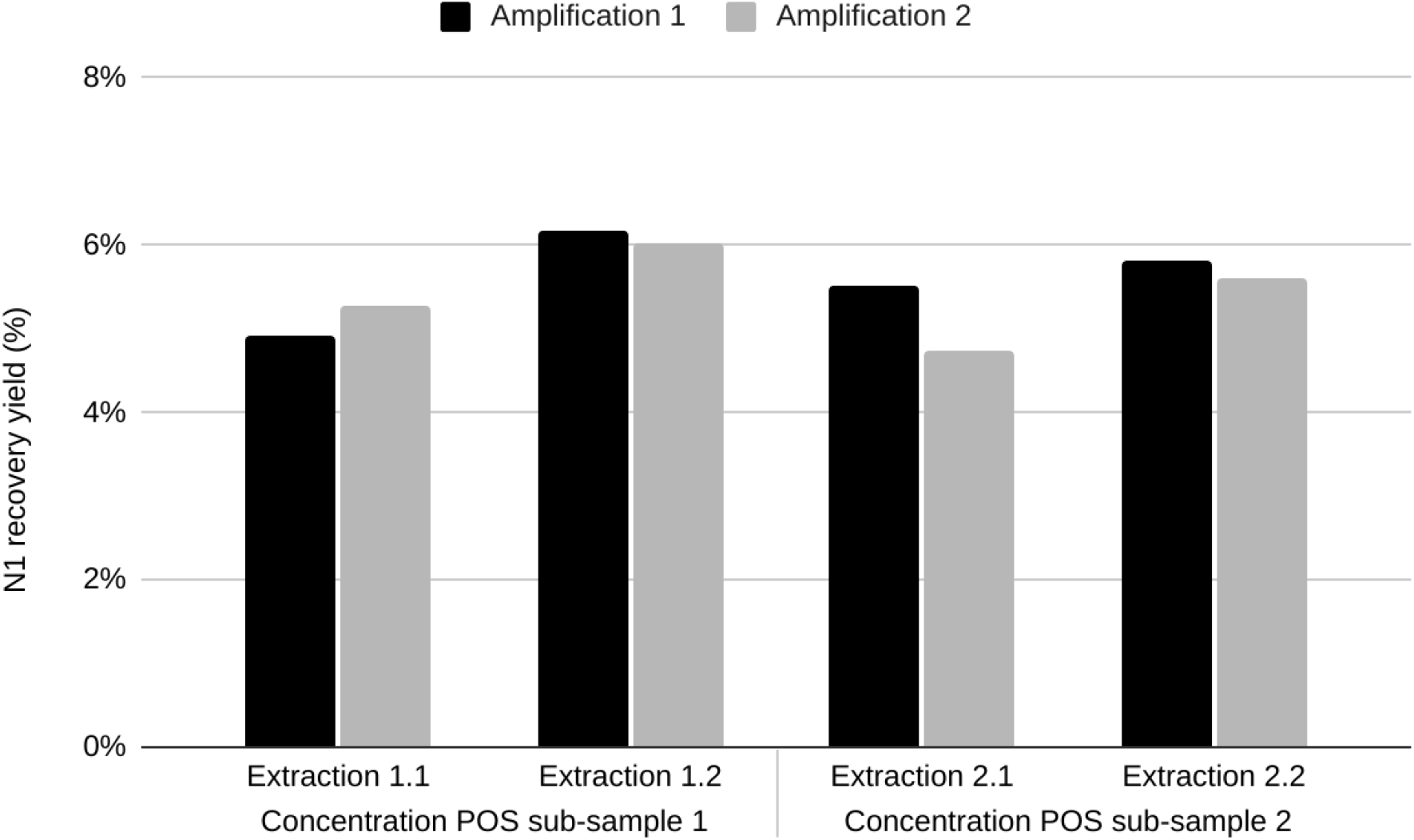
N1 recovery yield determination on a negative sample (NEG) after addition of heat inactivated SARS-CoV-2 (POS) in known quantity.

Several studies strongly suggest that centrifugal ultrafiltration units is a well-established and reliable method for WBE, describing acceptable recovery performance on model enveloped viruses (Bertrand et al., 2021; Jafferali et al., 2021; Ahmed et al., 2020b). In addition, 1 to 10% recovery yields are commonly observed in SARS-CoV-2 WBE (Gonzalez et al, 2020; Philo et al, 2021, Jafferaldi et al., 2021, D’aoust et al, 2021), and constitute the acceptable range of recovery according to ISO 15216-1:2017 standard. From that, the recovery yield determined in this study on heat-inactivated SARS-CoV-2 (5.5 +/-0.5%) was considered as satisfying. In contrast with other studies it is important to mention that results were not corrected by yield recovery.

In addition to the value of the recovery yield on heat-inactivated SARS-CoV-2, our experimental setup gave insight into the repeatability of concentration and extraction steps. As shown Figure 1, N1 recovery yield appeared statistically repeatable similar with regards to concentration replicates (with 5.6 +/-0.6 % for sub-sample/concentration 1 and 5.4 +/-0.5 % for sub-sample/concentration 2, t-test p-value >0.05). Extraction similarity was also observed as t-tests showed insignificant differences between replicates (with p-values >0.05) for both concentration sub-samples.

### Impact of solids separation on N1 titers on real wastewater sample

Most of the studies monitoring SARS-CoV-2 in wastewater process samples with a first solid separation step, in order to eliminate solid materials producing PCR inhibitors after genomic extraction. In the present work, we evaluated a preliminary solid separation step, namely clarification. Sample collected on September 7, 2020 on WWTP 7 was processed according to conventional direct-UF procedure as described in material and methods paragraph, but also submitted to pre-clarification (4000 *g*, 30 mn). In the last case, the pellet obtained after clarification was kept on ice before genomic extraction, whereas supernatant was subsequently submitted to UF.

The three samples (concentrate from direct UF, pellet from clarification and concentrate from subsequent UF) were submitted to the remaining standard procedure (*i*.*e*. triplicate extraction, from which two extractions were analysed in duplicate RT-qPCR).

The results obtained from this experiment showed higher N1 titers with direct UF than with pre-clarified UF method (Figure 2). N1 titers was 10^2.53 +/-0.16^ after direct UF, while it dropped to 10^1.49 +/-0.22^ after pre-clarification followed by supernatant UF (black bars on Figure 2).

**Figure 2:**
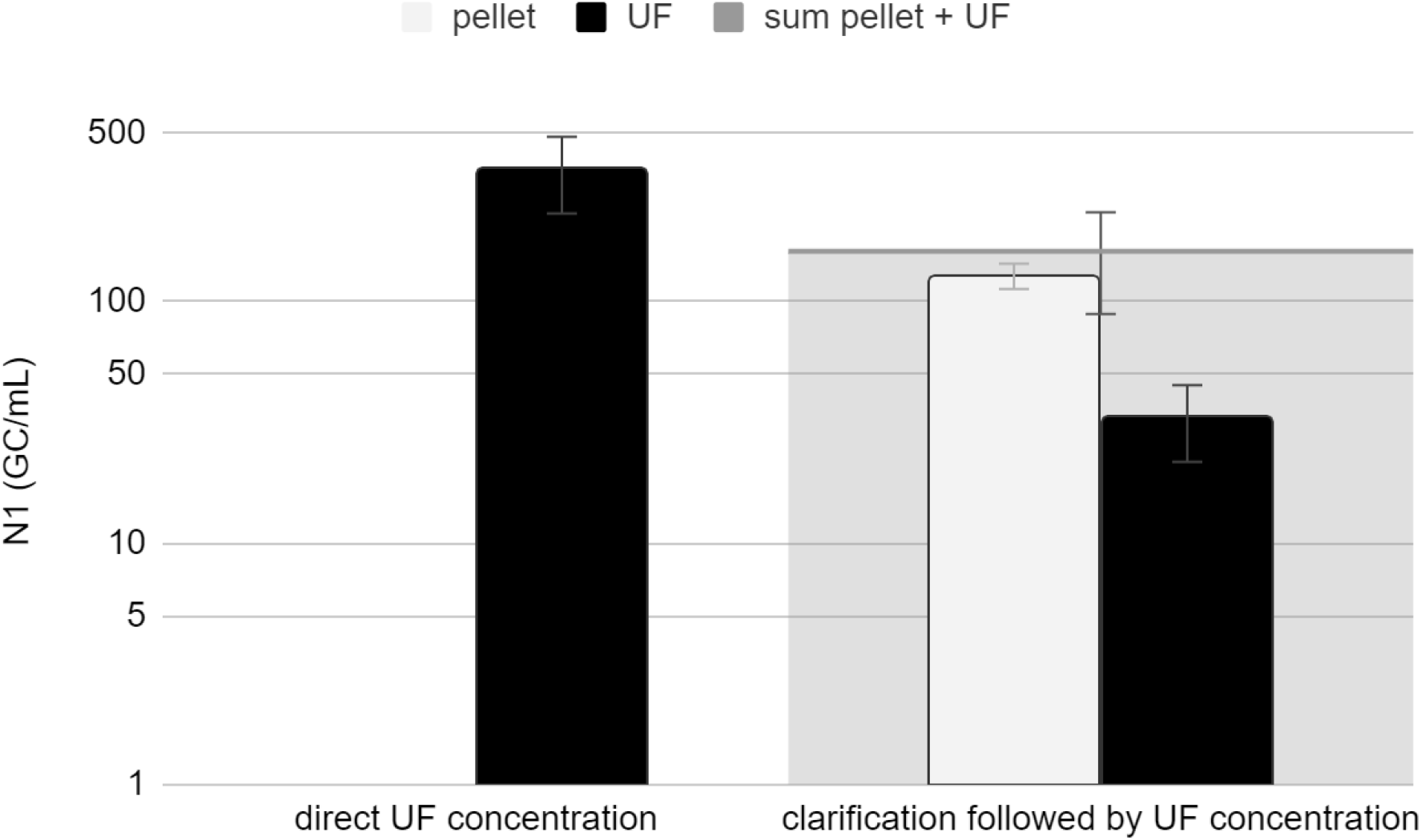
N1 titers in real wastewater sample, comparing direct UF concentration (1) method with pre-clarification followed by UF (2). UF concentrates (1&2) are depicted in black, pellet (2) in white and calculated pellet+UF (2) in light grey.

Interestingly, the pre-clarification pellet N1-titer was higher than the one observed in the pre-clarified supernatant UF concentrate (10^2.11+/-0.05^ against 10^1.49 +/-0.22^). In other terms, the pre-clarification pellet mainly contributed to N1 titer. Ultimately, combining pellet and preclarified UF concentrated wastewater yielded a lower N1 quantification than direct UF method (Figure 2, sum pellet + UF).

As an enveloped virus, SARS-CoV-2 is likely to be adsorbed onto the solid fraction of wastewater. Ye et al. (2016) have indeed reported higher solid-partitioning for enveloped viruses compared with non-enveloped viruses.

In this study, approximately 47% of the spiked MHV (Murine Hepatitis Virus) and 77% of the spiked ϕ6 were recovered in the centrate (liquid fraction) of the solids removed wastewater, suggesting that a relatively high fraction of the enveloped viruses (53% MHV and 23% ϕ6) were rapidly inactivated in the solids-removed wastewater (evaluated by cultivation).

Also in the present study, combining pellet with preclarified UF concentrated wastewater gave lower N1-quantification than direct UF, suggesting that viral genomic material was also rapidly affected by the solid removal step.

In addition, Ye et al. (2016) modelled solid-partitioning for two enveloped viruses (based on adsorption and inactivation kinetics) and predicted that up to 26% enveloped virus MHV adsorb to wastewater solids at equilibrium for a medium-strength municipal wastewaters with an average TSS value of 235 mg/L. While, SARS-CoV-2 detected by N1 target was found mainly (>50%) adsorbed to wastewater solids (superior quantification in pellet fraction than supernatant fraction) in the present study. It is important to note that this result was determined on only one wastewater sample and thus cannot be extrapolated to all wastewaters because, as also highlighted by Ye et al. (2016), wastewater solids concentration can vary widely. Consistently, TSS measured on this sample was higher than the one determined in Ye et al. (2016) study (with 366 mg-TSS/L) and yielded to higher virus fractionning on wastewater solids.

More recently, Kitamura et al. (2021) also evidenced solid fraction as an effective source for the detection of SARS-CoV-2 RNA in wastewater in comparison with concentrated supernatant. As their study focused on a low COVID-19 prevalence area (Japan), they revealed not only higher quantification but also higher detection rate on wastewater solid fraction compared to supernatant fraction. In recent studies, Fores et al. (2021) and Hokajarvi et al. (2021) also quantified up to 40% and more than 50% of SARS-CoV-2 RNA in pellet fraction of wastewater, respectively. Ultimately, as discussed by Chick et al. (2021) some authors have measured SARS-CoV-2 in primary sludge (Graham et al., 2021; Peccia et al., 2021; D’Aoust et al., 2021) and showed higher positive detection or quantification (up to 3,000 times) than in related wastewater samples in total accordance with the assumption of SARS-CoV-2 adsorption to wastewater solid fraction.

All together our results suggest that a non-negligible quantity of SARS-CoV-2 can be adsorbed on wastewater solid fraction. Again, this fractionning behavior is expected to be dependent on TSS content as suggested by Ye et al. (2016). Hence, our whole study was performed on total wastewater, performing direct UF. This direct protocol gave a benefit regarding quantification but also helped saving time and consumables.

However, it also strongly increased the risk of enzymatic inhibitors’ recovery. To ensure absence of inhibition, all 138 samples were appropriately diluted and controlled by SARS-CoV-2 synthetic RNA spiking. Spiking samples produced the expected concentration in all cases, with 10^3.38 +/-0.25^ N1 copies/µL and 10^3.50 +/-0.18^ N2 copies/µL in average in comparison with expected spike value of 10^3.60^, corresponding to less than 2 Cq value difference compared to the copy reference at the standard curve. Therefore, it was considered that there was no significant inhibition in all of our appropriately diluted samples.

### N1 and N2 targets response

Amongst the 138 analysed samples, 123 (89%) were positive with N1 probe while 114 (83%) were detected with N2 probe.

More precisely, amongst the 24 N2-negative samples, 14 (>50%) were also negative with the N1 probe, indicating undoubtedly-negative samples. One N1 negative-samples (<1% over the whole dataset) was positive with N2 probe, whereas 11 N2 negative-samples (8%) were detected with N1 probe.

In the context of wastewater networks, it is important to remind that SARS-CoV-2 is likely altered by the biotic and abiotic factors prevailing in such a complex ecosystem as suggested by La Rosa et al. (2020). This said, it is assumed that viruses can be partially degraded and so produce different concentrations regarding the tested biomarker, especially when the virus amount is low. Indeed, N1-N2 on-off difference concerns not only a very low amount of samples, but also samples depicting low- to moderate-SARS-CoV-2 RNA concentration (58 +/-50 UG/mL).

It is noteworthy to mention that despite small differences in terms of positivity frequency, N1 and N2 signals present a similar response trend in wastewater samples. Indeed, as illustrated on Figure 3, showing N1 and N2 quantification in the 138 wastewater samples plotted against sampling date, both biomarkers showed the same behavior when aggregating the 10 WWTPs.

**Figure 3:**
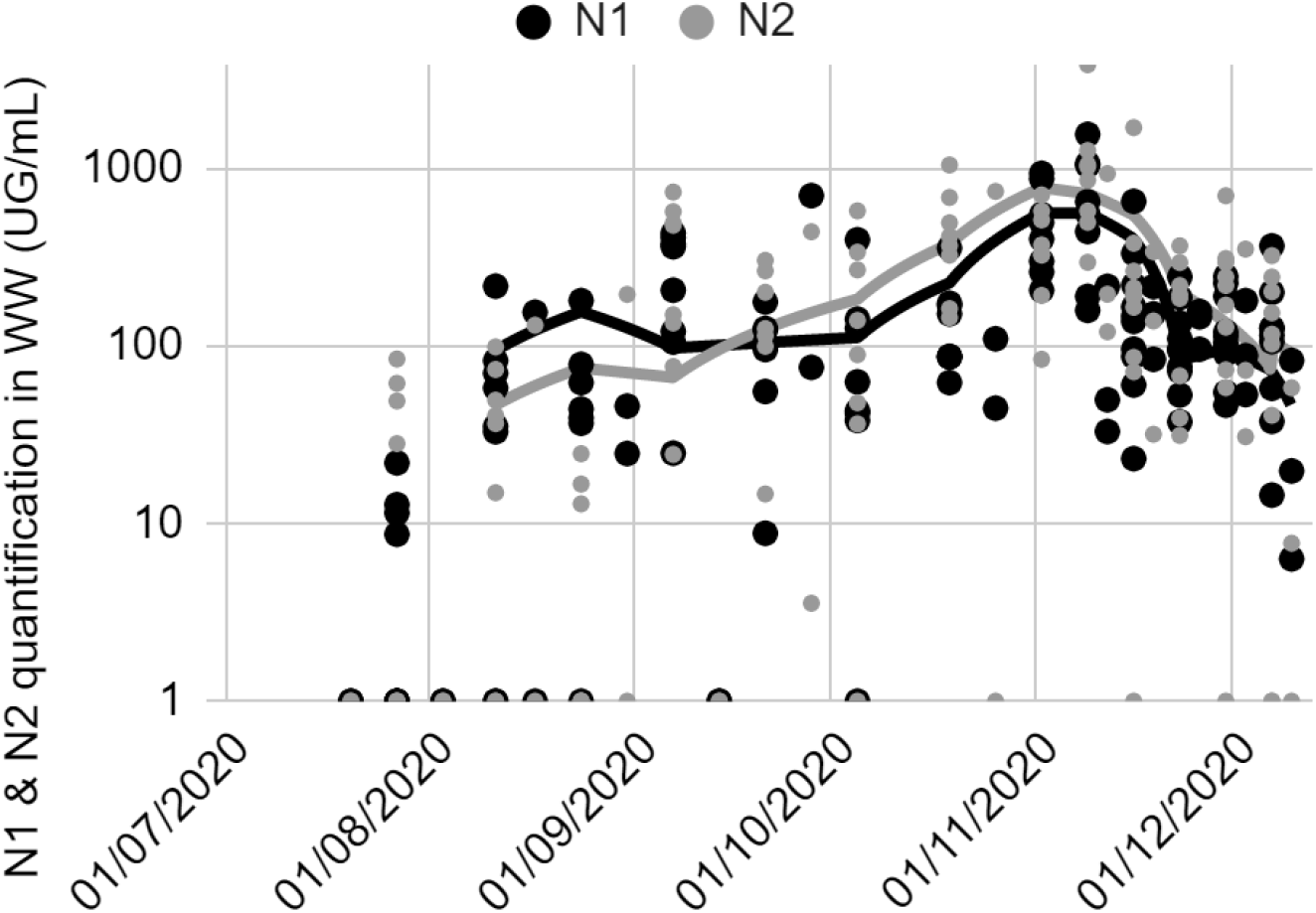
N1 and N2 quantification in the 138 wastewater-analysed samples along with sampling date. Smoothing curves correspond to centered moving averages (period 3).

These results strongly suggest that both probes can be used to follow SARS-CoV-2 prevalence in wastewater.

For more details, N1 and N2 quantification in 138 wastewater samples from 10 French sites between the end of July and the first fortnight of December revealed values in the ranges [10^0,3^ - 10^3,2^] UG/mL for N1 and [10^0,3^ - 10^3.6^] UG/mLfor N2, with 85% N1-values and 73% N2-values in the range [10 - 1,000] UG/mL. Interestingly, 45-50% measurements ranged in values between 100 and 1,000 UG/mL for both biomarkers, suggesting a good N1-N2 correlation. On the other hand, the range [10 - 100] was less represented in N2-values (24%) in comparison to N1-values (38%). Together with the highest proportion of N2-negative samples (than N1), this may reveal that N2 was less sensitive in the present study.

Such differences were already documented by other authors measuring SARS-CoV-2 RNA in wastewater samples as exemplified by the WRF interlaboratory work (Pecson et al., 2021), showing N1 to be more sensitive than N2 with up to 0.3 log difference. Also, contradictory results were observed by Gerrity et al. (2021) studying two WWTPs. N1 was more frequently recorded for one WWTP, whereas it was N2 for the second WWTP.

Nonetheless, N1 and N2 trends were likely similar and their correlation was tested using Spearman’s rank correlation method. Despite on-off N1-N2 behavior described before, Spearman’s correlation coefficient (ρ) was 0.79 (P-value <0.05), revealing a strong correlation strength between N1 and N2 measurements.

In addition, both quantifications showed variations with sampling date when monitoring SARS-CoV-2 in wastewater. To go deeper, these results have to be analyzed with regard to COVID-19 prevalence data, as well as sampling context (*e*.*g*. wet or dry weather, population connected). This work was done on N1 quantification, as this biomarker presented a better sensitivity in this study.

### WBE as a co-indicator of COVID-19 spread

Over the course of the 20-weeks study, 10 WWTP were monitored for SARS-CoV-2 RNA in the influent. These WWTP were all located in a different French administrative area (*Département*), and presented different plant characteristics. WWTP 1 & 2 were located in touristic areas, whereas the 8 others were related to metropolitan areas. As it can be seen on Table 1, monitored WWTP represented a relatively wide range of PE nominal treatment capacities, from 50,000 to 555,000 PE. This study focused on large WWTP as our aim was to compare wastewater-based SARS-CoV-2 RNA data to prevalence data, these last being collected and available at the level of “Département”.

**Table 1:**
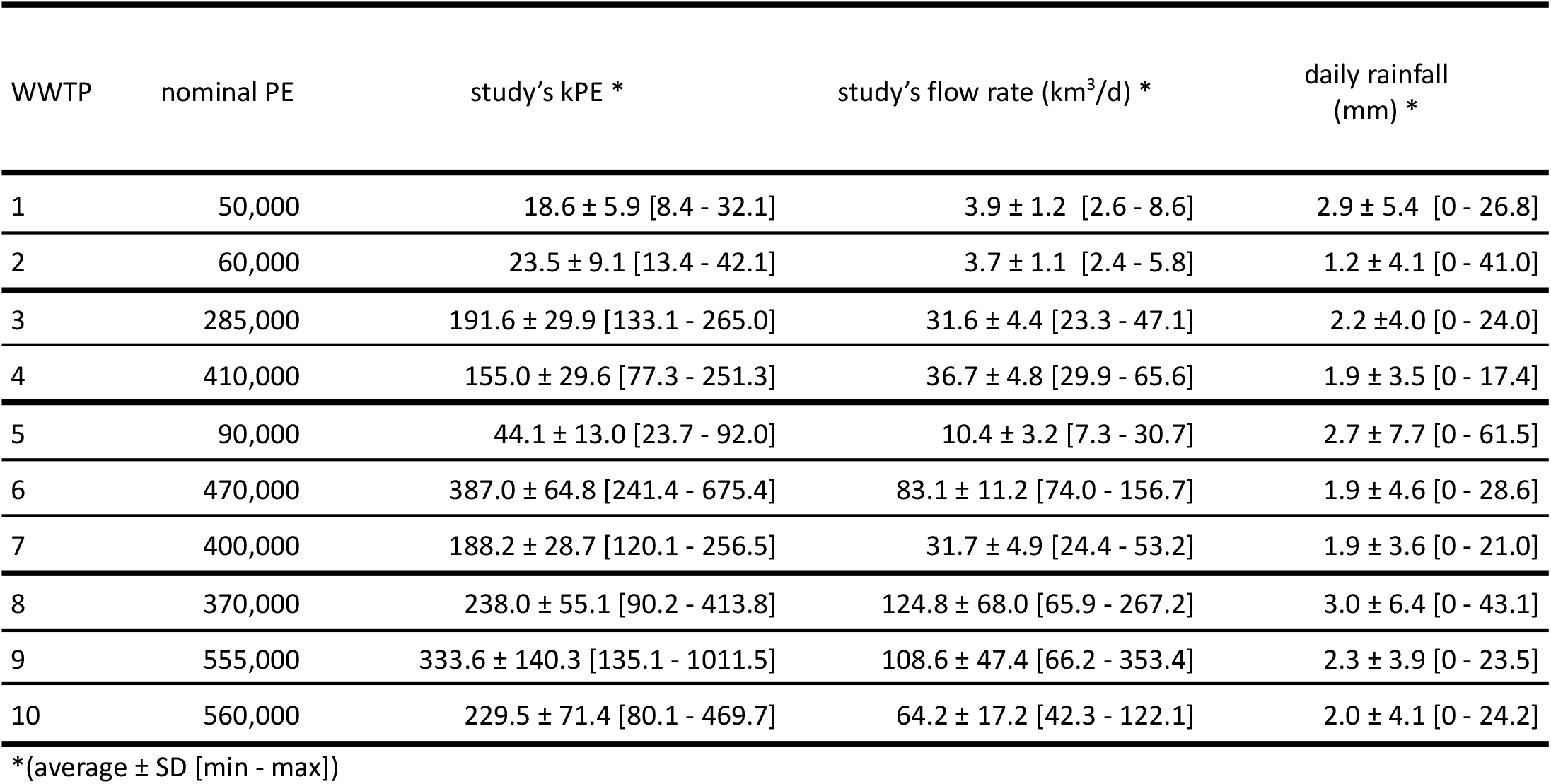
Wastewater treatment plants characteristics: organic loads, flow rate and rainfall

| WWTP | nominal PE | study's kPE * | study's flow rate (km<sup>3</sup>/d) * | daily rainfall (mm) * |
| --- | --- | --- | --- | --- |
| 1 | 50,000 | 18.6 ± 5.9 [8.4 - 32.1] | 3.9 ± 1.2 [2.6 - 8.6] | 2.9 ± 5.4 [0 - 26.8] |
| 2 | 60,000 | 23.5 ± 9.1 [13.4 - 42.1] | 3.7 ± 1.1 [2.4 - 5.8] | 1.2 ± 4.1 [0 - 41.0] |
| 3 | 285,000 | 191.6 ± 29.9 [133.1 - 265.0] | 31.6 ± 4.4 [23.3 - 47.1] | 2.2 ± 4.0 [0 - 24.0] |
| 4 | 410,000 | 155.0 ± 29.6 [77.3 - 251.3] | 36.7 ± 4.8 [29.9 - 65.6] | 1.9 ± 3.5 [0 - 17.4] |
| 5 | 90,000 | 44.1 ± 13.0 [23.7 - 92.0] | 10.4 ± 3.2 [7.3 - 30.7] | 2.7 ± 7.7 [0 - 61.5] |
| 6 | 470,000 | 387.0 ± 64.8 [241.4 - 675.4] | 83.1 ± 11.2 [74.0 - 156.7] | 1.9 ± 4.6 [0 - 28.6] |
| 7 | 400,000 | 188.2 ± 28.7 [120.1 - 256.5] | 31.7 ± 4.9 [24.4 - 53.2] | 1.9 ± 3.6 [0 - 21.0] |
| 8 | 370,000 | 238.0 ± 55.1 [90.2 - 413.8] | 124.8 ± 68.0 [65.9 - 267.2] | 3.0 ± 6.4 [0 - 43.1] |
| 9 | 555,000 | 333.6 ± 140.3 [135.1 - 1011.5] | 108.6 ± 47.4 [66.2 - 353.4] | 2.3 ± 3.9 [0 - 23.5] |
| 10 | 560,000 | 229.5 ± 71.4 [80.1 - 469.7] | 64.2 ± 17.2 [42.3 - 122.1] | 2.0 ± 4.1 [0 - 24.2] |

\*(average ± SD [min - max])

Daily organic load at the inlet of the WTTP on sampling days, the “study PE” in Table 1, were in the range [8,000 - 1,000,000] PE over the course of the study, all WWTP included. For WWTP 1, 2 and 5, daily organic loads were medium-range, with less than 100,000 PE in average, whereas the other ones were large-range with more than 100,000 PE in average. For each WWTP a relatively high PE variation (more than 10% of mean values) was observed along the study time course, especially for touristic areas (WWTP 1 & 2 depicting 30% PE variation). Indeed, the present study encompassed different seasons (summer and fall) corresponding to different population dynamics. As an example, during summer-time, population increases are expected in touristic areas and to lesser extent in urban areas WWTP influent flow rate was also varying along the study, with in all cases more than 10% variation against the mean. Importantly, flow rate variation can be attributed at least in part to PE variation, but also to rainfall, and whether sewers are combined or separate. In the case of combined sewer systems surface run-off during rain events are collected in the same pipe as wastewater, both increasing WWTP influent flow rate and diluting it during high rainfall events. It can be noted from Table 1 that WWTP 8, 9 and 10 presented dramatic variations in flow rate, with more than 25% of mean values.

Regarding SARS-CoV-2 RNA monitoring, all samples were collected from flow-driven 24h auto-sampler, producing composite samples that were transported to the laboratory on ice and analysed within maximum 4 days after collection.

Sampling frequency was first twice a month (from the end of July to the beginning of November 2020) and then increased to once a week for 7 WWTPs and twice a week for 3 WWTPs until December 2020.

For each ten “Départements”, COVID-19 prevalence data were collected from the open data portal of the French National Health Agency and compared to SARS-CoV-2 N1 quantification in wastewater.

As presented in Figure 4, both measures presented the same trend with time when gathering the 10 WWTPs. A relatively continuous increase was observed from late July to the first days of November, followed by a first sharp then slow decrease until mid-December. Consistently, a second unlock was implemented in France from October 30 until December 15. This governmental decision appeared in relation with the drop of both disease incidence rate and wastewater monitoring.

**Figure 4:**
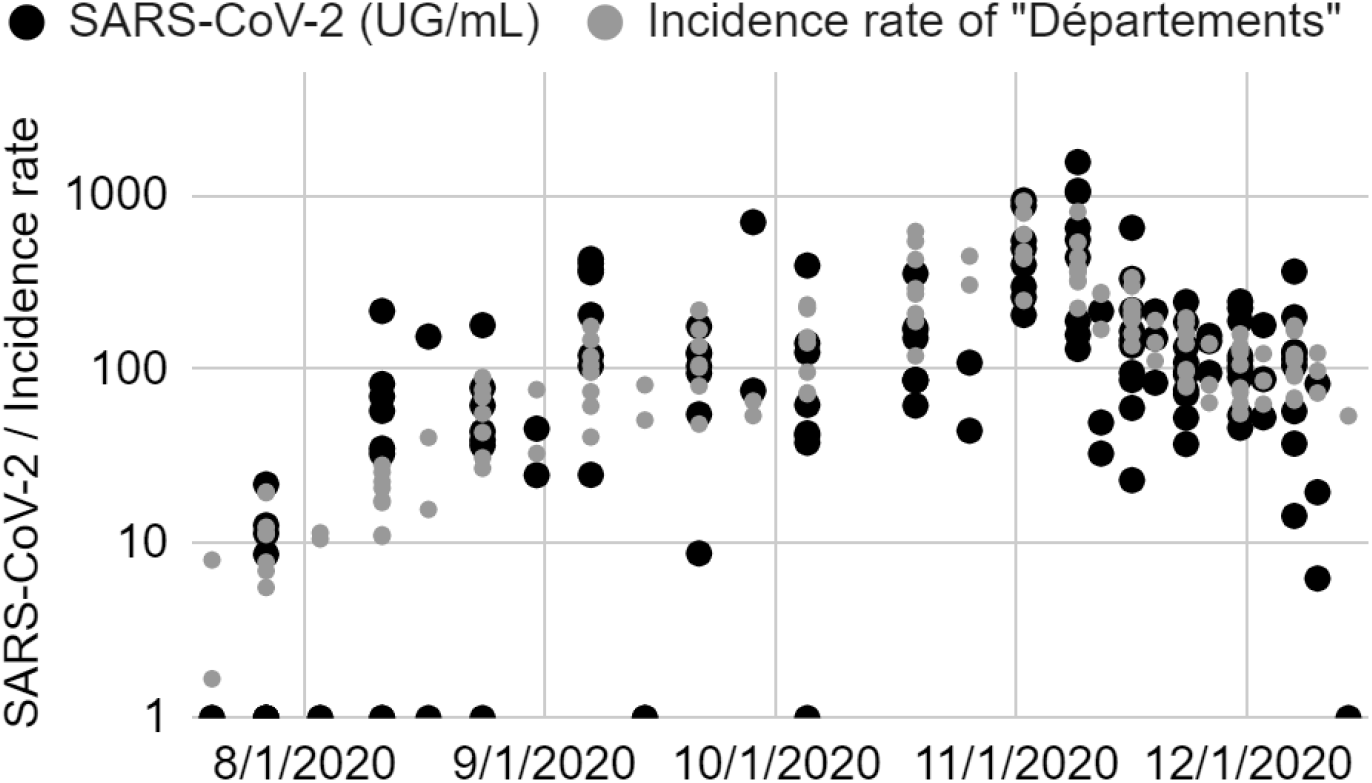
N1 quantification in the 138 wastewater analysed samples in comparison with incidence rate of “Départements” along with sampling date.

Spearman’s correlation between wastewater SARS-CoV-2 RNA and COVID-19 prevalence at the scale of “Département” was evaluated and revealed a medium-strong strength (ρ = 0.66) when looking at the 10 WWTPs altogether. This result strongly supports SARS-CoV-2 RNA monitoring in wastewater as a good co-indicator of epidemic spread, together with incidence rate.

For a more detailed vision, N1-SARS-CoV-2 measure in wastewater was plotted together with incidence rate of “Département” for each WWTPs against time (Figure 5).

**Figure 5:**
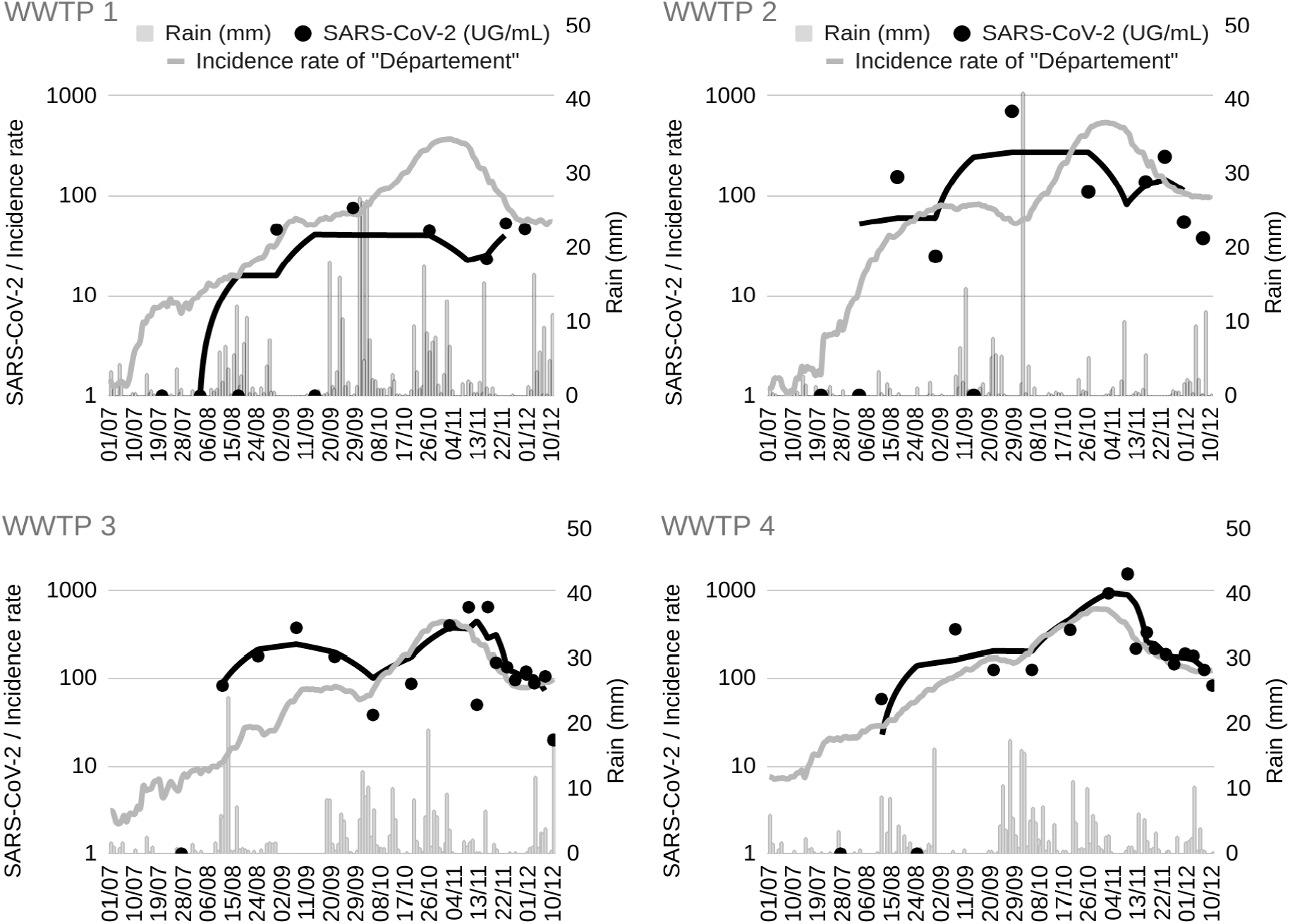
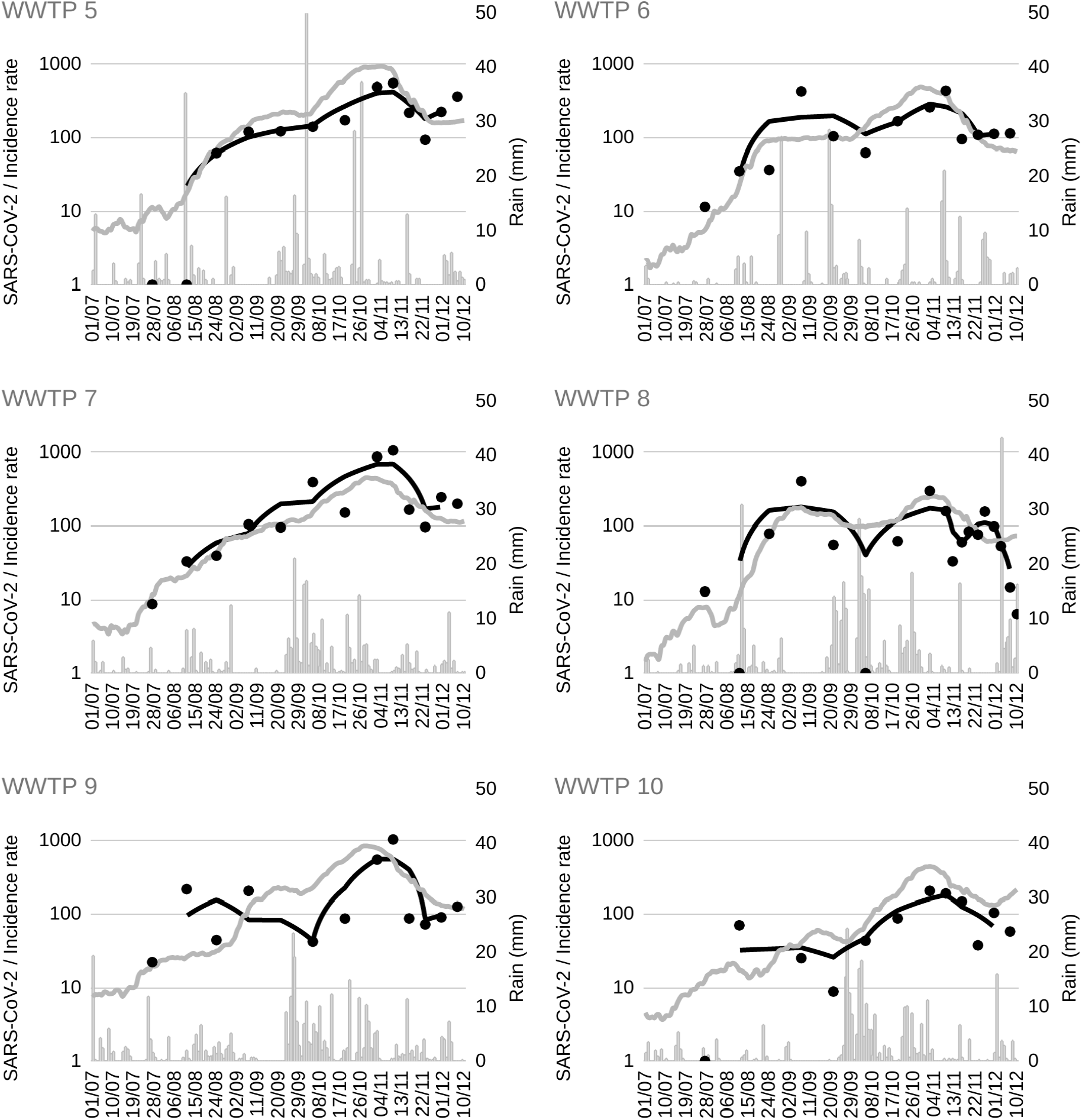
Raw N1-SARS-CoV-2 quantification (SARS-CoV-2, black circles and black moving average), incidence rate of “Département” (light grey curve) and rain (light grey bars) along with date for the 10 studied WWTPs.

A quick look at these 10 graphs shows that SARS-CoV-2 RNA was detected in all WWTPs. As the time course of the study encompassed a globally alarming sanitary situation and a lockdown event, it was not surprising but consistent to find SARS-CoV-2 genomic traces in all studied locations.

WWTP 1 and WWTP 2, which are located in touristic areas, presented rather perturbed SARS-CoV-2 RNA measurements profiles in wastewater. Indeed, from late July to late September both WWTP signals showed discrepancies with time and on/off behaviors. One main explanation for such strong variations is population dynamic in touristic area during summer 2020, inducing variability in the sampling population evaluated by wastewater collection. Also it is noteworthy to mention that these WWTPs accounted for 50,000-60,000 PE whereas French “*Départements”* represent about 1 to 2 million people and thus it is unclear how data representing such different spatial units can be compared.

Nonetheless, from the end of summer it is clear that coronavirus RNA was quantified in relatively high amounts, consistently with the epidemic spread observed throughout the incidence rate.

For WWTP 3 to 10, epidemiological and wastewater signals were visually following similar trends. This result suggests that metropolitan areas monitoring with WBE fits better with epidemiological data than for touristic sites when population movements are permitted (*i*.*e*. during summer time). Also as the incidence rate of “Département” reflects all the positive biological tests collected on an about 1 to 2 million population scale, it is consistent that it fits better with WBE data collected on medium-high sized WWTP than smaller ones. Comparison of WBE data with local incidence rate is highly recommended but for statistical secrecy reasons such data cannot be open-accessible in France.

Thus, Spearman’s correlations between WBE data and incidence rate of “Département” were evaluated on the 10 WWTPs. As shown in Table 2, Spearman’s correlation coefficients were between 0.32 and 0.82 over the ten WWTPs. Assuming that 0-0.3 correlation coefficient stands for low correlation, 0.3-0.5 for medium and 0.5-1 for strong correlation (Hinkle et al., 2003), 4 and 6 WWTPs presented medium and strong WBE data correlation with incidence rate of “Département”, respectively. To be more precise, 6 cases depicted significant p-values (< 0.05) regarding spearman correlation. Consistently, non-significant p-values (> 0.05) were mainly related to low correlation strength.

**Table 2:** Correlation analysis between SARS-CoV-2 WBE data (raw and load data) and COVID-19 incidence rate of “Département” (Spearman’s correlation coefficients ρ, blank columns) and linear regression between inlet flow rate and rain (slope α and R^2^, grey columns) for the ten studied WWTP.

| WWTP | $\rho$<br>raw SARS-CoV-2<br>vs. incidence rate<br>(log10) | slope ($\alpha$)<br>% norm. inlet flow<br>rate variation vs.<br>rain | $R^2$<br>% WW flow rate<br>variation vs.<br>rain | $\rho$<br>contextualized<br>SARS-CoV-2 load/PE vs.<br>incidence rate (log10) |
| --- | --- | --- | --- | --- |
| 1 | 0.58* | 7.0 | **0.51** | **0.87** |
| 2 | 0.41* | NA | NA | 0.36* |
| 3 | 0.32* | 3.1 | 0.25 | 0.30* |
| 4 | **0.72** | 3.5 | 0.49 | **0.70** |
| 5 | **0.78** | 5.3 | **0.57** | **0.81** |
| 6 | **0.58** | 2.9 | **0.55** | **0.71** |
| 7 | **0.81** | 4.8 | **0.61** | **0.86** |
| 8 | 0.35 | 18.8 | **0.72** | **0.54** |
| 9 | 0.32* | 14.5 | **0.78** | **0.66** |
| 10 | **0.82** | 9.4 | **0.79** | **0.85** |

\*stands for p-values>0.05, no star means p-value<0.05

Other studies already reported such consistency between WBE and epidemiological data (Medema et al. 2020a, Agrawal et al. 2021, Hata et al. 2020, D’Aoust et al. 2021). Medema et al. (2020a) and Agrawal et al. (2021) evidenced strong correlations (0.6 - 0.8) and highlighted WBE data dispersion as well as local testing and tracing policies as variability purveyors. On the contrary, Hata et al. (2020) and D’Aoust et al. (2021) recorded low-to-medium correlation strengths (0.3 - 0.5), the last advancing the need for normalization to reduce noise.

Despite some noise during summer time, our results were consistent with previous studies and impressively supporting WBE as a powerful tool for epidemiological surveillance. It is important to recall that COVID-19 incidence rates were reported daily as 7-day sums to take into account data reporting delays. In this situation, it is unclear whether it is possible or not to compare time series on a daily basis. In addition, sampling frequency being twice a month from late July to the beginning of November, time shifts were not considered in this study.

Interestingly for WWTPs 3, 6, 8 and 9 WW SARS-CoV-2 RNA quantification appeared highly affected by high rainfall events. It was particularly remarkable for WWTP 8 results on October 5 and December 7 and 10, when rain not only diluted the signal but in this case turned it off. Obviously, it doesn’t mean that the virus suddenly stopped circulating in the population.

To go deeper and based on the visual observation of high rainfall events co-occurrence with WBE data drop, we studied the link between rainfall and wastewater flow rate during the course of the study over the 10 WWTP. For that we looked at linear regressions between 7-days average data. Indeed, it is well known that rainfall can impact flow rate with a lag time, depending on runoff dynamic, soil drying as well as sewer network characteristics. Also, in order to compare WWTPs (presenting a wide average flow rate range, Table 1) each flow rate time measures were normalized by the corresponding WWTP averaged flow rate in dry weather conditions, knowing that dry weather conditions were determined as a period of 7 days without rain. This calculation (reported to 100%) produced a normalized flow rate that can be interpreted as flow rate variation against dry weather conditions.

Linear regression modelling was performed on dry-weather normalized flow rate (7-days averaged) against rainfalls (7-days averaged). Except for WWTP2 where linear model was not applicable (residuals presenting a coma pattern), all WWTPs can be modeled with linear regressions with slope and regression coefficient p-values <0.05 and roughly normal and homoscedastic residuals.

For ‘touristic’ WWTPs 1 & 2, contrasting results were observed, as WWTP 2 didn’t satisfy modeling conditions and WWTP 1 presented a medium-to-strong correlation coefficient (>0.5, Table 2). Importantly, for WWTP 2 flow rate variation was not explained by rainfall, suggesting the presence of another factor influencing flow rate. Interestingly, a strong flow rate increase that co-occurred with a strong PE increase was observed during summer time (data not shown), highly suggesting population influx being a preponderant predictor to flow rate variation for this touristic WWTP during the studied period.

By contrast, WWTP 1 flow rate variation can be at least in part predicted by rainfalls with a relatively medium-to-high gradient (α = 7.0 % flow-rate variation/rainfall mm), meaning that rainfall may have a relatively high effect on flow rate variation.

For metropolitan WWTPs 3 to 10, dry weather normalized flow rate variation linear relationship with rainfalls revealed 3 behaviors classified as low-, medium- and high-strength. WWTP 3 & 4 presented low-strength correlation, with low correlation coefficient (0.25 and 0.49 for WWTP 3 & 4, respectively) and relatively low gradient (α = 3.1 and 3.5 for WWTP 3 & 4, respectively).

WWTP 5, 6 & 7 correlations showed medium-strength with medium-to-strong correlation coefficients (0.57, 0.55 and 0.61, respectively) and low-to-medium gradients (α = 5.3, 2.9 and 4.8, respectively).

And WWTPs 8, 9 & 10 depicted strong correlation strength revealed by strong correlation coefficients (0.72, 0.78 and 0.79, respectively) and high gradients (α = 18.8, 14.5 and 9.4, respectively).

From this, it is clear that the link between rainfall and flow rate is dramatically different between WWTPs. Simple linear regressions can obviously not reflect nor demonstrate the causal relationship between variables, but based on the knowledge of sewer systems structuration that presents combined and/or separate sub-systems, these results suggest WWTPs 8, 9 & 10 (and to a lesser extent WWTPs 5, 6 & 7) as to be more likely mainly presenting combined sewers system in comparison with others studied WWTPs.

In terms of quantification this would imply a potential dilution effect of rainfall more or less important depending on WWTPs. In an attempt to tackle this phenomenon, we impacted SARS-CoV-2 RNA quantifications by the daily flow rate/averaged dry conditions flow rate ratio, implying a calculated increase of SARS-CoV-2 RNA quantification values when flow rate is higher than mean flow rate. Also SARS-CoV-2 RNA concentrations were converted into viral loads that were reported to PE, knowing that PE was defined as 60g-BOD_5_/day. This calculation produced corrected values thereafter referred to as contextualized N1-SARS-CoV-2 load.

To observe the effect of such an approach on the ten WWTP, contextualized N1-SARS-CoV-2 loads were plotted together with raw SARS-CoV-2 quantification as well as incidence rate of the *Département* and daily rainfall (Figure 6).

**Figure 6:**
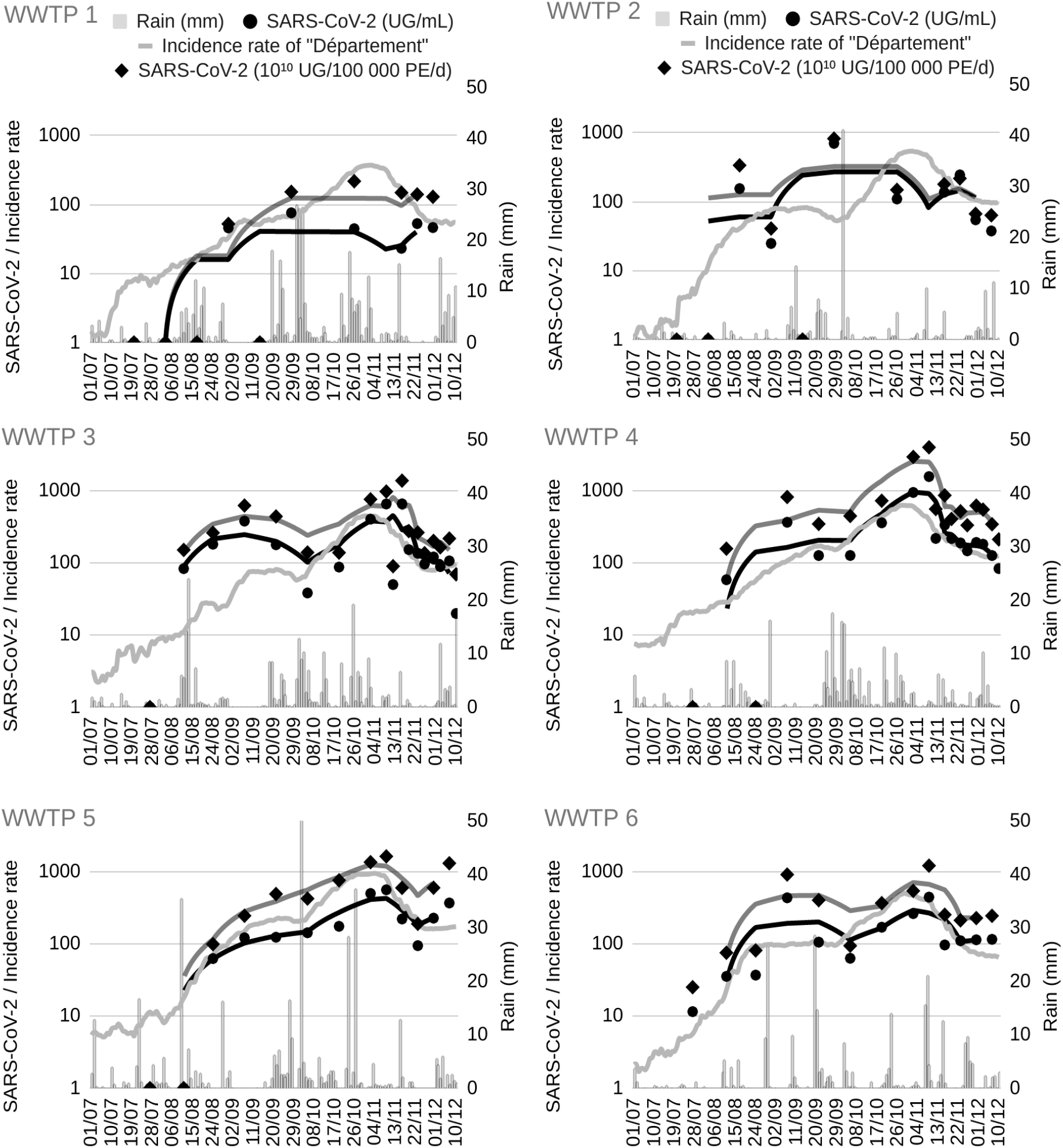
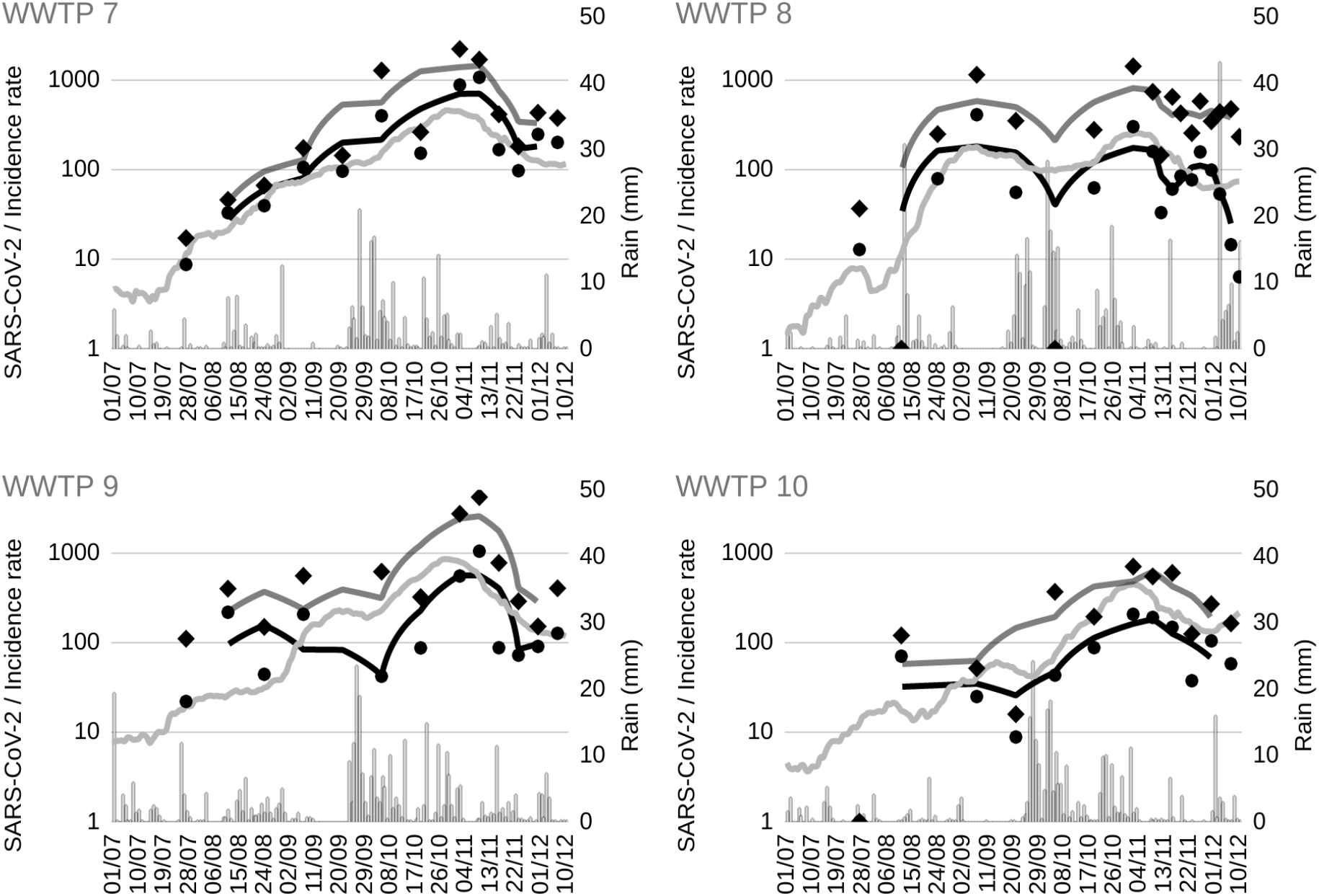
Raw N1-SARS-CoV-2 quantification and contextualized N1-SARS-CoV-2 load (SARS-CoV-2 UG/mL, black circles and black moving average and SARS-CoV-2 10^10^ UG/ 100 000 PE/d, black diamonds and dark grey moving average), incidence rate of “Département” (light grey curve) and rain (light grey bars) along with date for the 10 studied WWTPs.

As expected, for the low strength rainfalls/influent flow rate variation correlation WWTPs (2, 3 &4), dynamics were poorly impacted by the proposed calculation. Indeed, as it can be seen on Figure 6 (WWTP 2, 3 &4), contextualized SARS-CoV-2 RNA load followed the same trend as raw SARS-CoV-2 RNA quantification. Consistently, Spearman’s correlation between contextualized load and prevalence of the *Département* didn’t show any improvement in comparison with raw quantification SARS-CoV-2 RNA data (Table 2).

But for medium- and high-strength rainfalls/influent flow rate variation correlation (WWTPs 1, 5, 6 & 7 and 8, 9 & 10, respectively), there was a visually noticeable impact on contextualized loads when compared to raw quantification on log scale (Figure 6). This impact was consistently more pronounced as rain/flow rate variation correlation was high. Particularly, for WWTP 8, the last part of the two curves differed dramatically, with raw data dropping and contextualized load flattening during the first fortnight of December.

Regarding WWTP 9, relatively high rainfall occurring at the beginning of October were consistently taken into account in contextualized load data, producing a stable signal from the beginning of the study until this autumnal high precipitation period, in contrast with decreasing trend observed on raw quantification.

WWTP 10 also presented rainfall events in early Autumn and in this case, contextualization induced a time-shift - contextualized data being in advance compared to raw quantification data.

Interestingly, Spearman’s correlation between contextualized loads and the disease prevalence in the *Département*, showed an improvement for all studied WWTPs presenting the medium- to high-strength rainfalls/influent flow rate variation correlation (Table 2). Overall for these 7 WWTPs (1, 5, 6, 7, 8, 9 & 10) Spearman’s correlation coefficients achieved values > 0.5 and in many cases as high 0.8. This result strongly suggests that rain impact on SARS-CoV-2 RNA quantification in combined sewer systems can be corrected with appropriate data collection, producing highly valuable information for pandemic monitoring. As raw SARS-CoV-2 RNA quantification data already presented a good fit with disease prevalence data, it is important to say that this finding does not imply a crucial need for data correction when monitoring SARS-CoV-2 RNA in wastewater. But rather it highlights that these data ultimately need to be contextualized and analyzed by people who have a local and territorial knowledge of sewer system structuration, as local authorities or WWTP operators.

Previous authors have already suggested that sampling context (*e*.*g*. wet or dry weather, population connected) may impact WBE quantitative results but to our best actual knowledge, our work constitutes the first example confirming it on medium-term SARS-CoV-2 RNA monitoring data from a meaningful number of WWTP spread throughout different areas.

## Conclusions

A reliable protocol for the quantification of SARS-CoV-2 RNA in wastewater was developed and applied to samples coming from a broad range of WWTP. This protocol combining ultrafiltration - RNA extraction - RT-qPCR, was qualified using heat-inactivated SARS-CoV-2 and we obtained a recovery yield of 5.5 +/-0.5%. Moreover, direct UF on both liquid and solid fractions from wastewater gave satisfying results regarding quantification. As for N1 and N2 target responses, their trends were similar with a strong correlation. N1 was picked, as it appeared to be the most sensitive biomarker here. Ultimately, the protocol is operational, quick and easy to implement and benefits from a specific and sensitive PCR.

SARS-CoV-2 RNA monitoring in wastewater appeared to be consistent with the COVID-19 prevalence data. Qualitatively, wastewater reflected well the epidemic dynamics in France, especially its peak preceding the second lockdown and its subsequent decline. Our study confirmed a correlation between the incidence rate and the presence of SARS-CoV-2 RNA in wastewater, shown by other authors. In our case, this correlation can be qualified as strong to medium. The consistency with traditional epidemiological data can be observed globally, for all ten WWTP together. Depending on the WWTP, the correlation was more or less strong, due to the effects of wet weather and population variation. Therefore, to avoid those effects, we looked further than the daily average concentrations of SARS-CoV-2 RNA and computed the contextualized daily loads per inhabitant. This possible impact of sampling context on WBE has been suggested by several authors. Nevertheless, to our best knowledge, our study constitutes the first example confirming its relevance on medium-term SARS-CoV-2 monitoring data from a meaningful number of WWTP spread throughout different regions and serving different populations. Accounting for the sampling context is thus fundamental to properly interpret SARS-CoV-2 RNA monitoring in sewers.

## Data Availability

All data and methods are reported in the manuscript

## Credit authorship contribution statement

**Adèle Lazuka:** Conceptualization, Formal Analysis, Methodology, Investigation, Visualization, Writing Original draft

**Anne-Sophie Lepeupl**e: Study design, supervision, Review

**Charlotte Arnal:** Investigation, Formal analysis, Writing, Review

**Emmanuel Soyeux**: Investigation, Formal analysis, Writing, Review

**Mickael Sampson**: Methodology, Analysis

**Yannick Deleuze**: Formal Analysis, Visualization, Review

**Stanislas Pouradier Duteil:** Resources and Review

**Sébastien Lacroix**: Conceptualization, Study design, Methodology, Supervision, Writing Review and Editing

All authors discussed the results and commented on the manuscript

## Acknowledgements

We would like to acknowledge Estelle Gonidec, Camille Negre, Florian Gauthier-Fournier and Matthieu Cormenier for technical support. Sophie Vaudran is also gratefully acknowledged for crucial help with logistics. The authors further acknowledge the effort made by Christelle Pagotto and Sandra Fuentes from Veolia Eau France Technical Direction for assisting sample collection and Veolia Eau France operators for sampling. Oliver Keserue kindly accepted to review the english writing.

